# Longitudinal changes in age and race of patients with SARS-CoV-2 in a multi-hospital health system

**DOI:** 10.1101/2021.08.16.21262016

**Authors:** Ian J. Barbash, Lee H. Harrison, Jana L. Jacobs, Faraaz Ali Shah, Tomeka L. Suber, Kevin L. Quinn, Oscar C. Marroquin, Mark T. Gladwin, Rachel E. Sackrowitz, Alison Morris

## Abstract

**Background:** The COVID-19 pandemic continues to affect the United States and the world. Media reports have suggested that the wave of the alpha variant in the Spring of 2021 in the US caused more cases among younger patients and racial and ethnic subgroups.

**Approach:** We analyzed electronic health record data from a multihospital health system to test whether younger patients accounted for more cases and more severe disease, and whether racial disparities are widening. We compared demographics, patient characteristics, and hospitalization variables for patients admitted from November 2020 through January 2021 to those admitted in March and April 2021.

**Results:** We analyzed data for 37, 502 unique inpatients and outpatients at 21 hospitals from November 1, 2020 to April 30, 2021. Compared to patients from November through January, those with positive tests in March and April were younger and less likely to die. Among patients under age 50, those with positive tests in March and April were three times as likely to be hospitalized and twice as likely to require ICU admission or mechanical ventilation. Individuals identified as Black represented a greater proportion of cases and hospitalizations in March and April as compared to November through January.

**Conclusions:** We found that relative COVID-19 hospitalization rates for younger individuals and individuals identified as Black were rising over time. These findings have important implications for ongoing public health measures to mitigate the impact of the pandemic.

## INTRODUCTION

The pandemic of novel coronavirus disease 2019 (COVID-19) caused more than 600,000 deaths by June 2021 in the United States alone (1). From the outset, the pandemic has disproportionately affected essential workers, individuals living in economically disadvantaged neighborhoods, and Black and Hispanic communities (2). The widespread availability of highly effective vaccines holds promise, but infections and deaths from the disease continue. Despite falling cases this year, media reports indicate that younger individuals and those identified as Black now account for a greater proportion of infections and deaths than in earlier phases of the pandemic—some of the same groups in which vaccination rates remain lower (3, 4). This dynamic is particularly concerning in light of the continued emergence of novel SARS-CoV-2 variants (5). We examined data from a multi-hospital health system to determine whether younger patients now account for more cases and more severe disease, and whether racial disparities are widening.

## METHODS

We evaluated data from 21 hospitals within the UPMC Health System between November 1, 2020 and April 30, 2021. We included inpatients aged 18 or older who had acute infection with SARS-CoV-2 based on the results of internal and external laboratory tests. We used UPMC’s Clinical Data Warehouse (CDW) to obtain data on demographics, selected comorbidities, inpatient medication utilization (remdesivir and corticosteroids), admission to an intensive care unit (ICU), requirement for invasive mechanical ventilation, and inpatient mortality. In the event of multiple eligible hospitalizations, we included only the first hospitalization. We considered interhospital transfers as a single episode of care. All inpatients had a minimum of one month of follow-up data through May 31, 2021. We also obtained data from the CDW for the same time period on outpatients age 18 or older with positive SARS-CoV-2 test results to characterize the population at risk for hospitalization.

We compared patients admitted between November 1, 2020 and January 31, 2021 to patients admitted between March 1, 2021 and April 30, 2021, excluding February to reduce cross-period contamination. We analyzed differences in age, race, comorbidities, requirement for mechanical ventilation, and hospital and ICU admission rates between the two periods, using standard statistics. We performed analyses in which the denominator was either 1) patients with inpatient or outpatient positive SARS-CoV-2 tests or 2) patients admitted with COVID-19. We analyzed the overall cohort as well as the subgroup of patients under age 50. We also attempted SARS-CoV-2 whole genome sequencing (WGS) directly from residual endotracheal aspirate specimens from five younger patients who experienced a rapid clinical decline during admission in March and April, 2021 (6).

## RESULTS

We identified 29,728 unique patients with outpatient positive tests who were not admitted and 7,774 unique patients admitted with COVID-19. Compared to patients identified in November through January, those from March and April were more likely to be of Black race (17.5 vs. 7.5%, p<0.001) and more likely to be hospitalized (28.5% vs. 19.5%, p<0.001), but similarly likely to require ICU admission or mechanical ventilation (Table 1). Among hospitalized patients, those admitted in March and April were younger (median age 64 vs. 71, p<0.001), more likely to be identified as Black (20.2% vs. 11.3%, p<0.001), less likely to have been admitted to an ICU or mechanically ventilated, and less likely to die (Table 2).

**Table 1.** Characteristics of inpatients and outpatients with SARS-CoV-2 infection by period

|  |  |  |  |  |
| --- | --- | --- | --- | --- |
| All patients |  | Nov-Jan | Mar-Apr | p-value |
|  |  | N= 32307 | N= 5195 |  |
| Age, median (IQR) |  | 54 (37, 68) | 53 (38, 65) | <0.001 |
| Race | Black | 2435 (7.5%) | 909 (17.5%) | <0.001 |
|  | White | 28525 (88.3%) | 4096 (78.8%) |  |
|  | Other | 1347 (4.2%) | 190 (3.7%) |  |
| Hypertension |  | 9420 (29.2%) | 1536 (29.6%) | 0.55 |
| Diabetes |  | 5134 (15.9%) | 866 (16.7%) | 0.16 |
| COPD |  | 3690 (11.4%) | 689 (13.3%) | <0.001 |
| Hospitalized |  | 6294 (19.5%) | 1480 (28.5%) | <0.001 |
| ICU |  | 1450 (4.5%) | 258 (5.0%) | 0.13 |
| Mechanical Ventilation |  | 756 (2.3%) | 112 (2.2%) | 0.41 |
| Hospital Mortality |  | 1265 (3.9%) | 64 (1.2%) | <0.001 |
| Patients under age 50 |  |  |  |  |
|  |  | N= 13676 | N= 2267 |  |
| Age, median (IQR) |  | 34.3 (26.5, 42.5) | 35 (26.8, 42.9) | 0.23 |
| Race | Black | 1203 (8.8%) | 499 (22.0%) | <0.001 |
|  | White | 11638 (85.1%) | 1660 (73.2%) |  |
|  | Other | 835 (6.1%) | 108 (4.8%) |  |
| Hypertension |  | 1619 (11.8%) | 311 (13.7%) | 0.011 |
| Diabetes |  | 668 (4.9%) | 134 (5.9%) | 0.038 |
| COPD |  | 848 (6.2%) | 172 (7.6%) | 0.012 |
| Hospitalized |  | 622 (4.5%) | 283 (12.5%) | <0.001 |
| ICU |  | 141 (1.0%) | 49 (2.2%) | <0.001 |
| Mechanical Ventilation |  | 79 (0.6%) | 25 (1.1%) | 0.004 |
| Hospital Mortality |  | 28 (0.2%) | 3 (0.1%) | 0.47 |
| Abbreviations: ED=Emergency Department; COPD=Chronic Obstructive Pulmonary Disease; LOS=Length of Stay; ICU=Intensive Care Unit; IQR=Interquartile Range |  |  |  |  |
| Statistical analysis: categorical variables are displayed as N(%) and analyzed with chi-squared tests. Continuous variables are displayed as median (IQR) and analyzed using Wilcoxon Rank-Sum tests. |  |  |  |  |

**Table 2.** Characteristics of inpatients with SARS-CoV-2 infection by period

|  |  |  |  |  |
| --- | --- | --- | --- | --- |
| All patients |  | Nov-Jan | Mar-Apr | p-value |
|  |  | N=6294 | N=1480 |  |
| Age, median (IQR) |  | 71 (61, 80) | 64 (53, 74) | <0.001 |
| Race | Black | 709 (11.3%) | 299 (20.2%) | <0.001 |
|  | White | 5365 (85.2%) | 1139 (77.0%) |  |
|  | Other | 220 (3.5%) | 42 (2.8%) |  |
| Hypertension |  | 2757 (43.8%) | 622 (42.0%) | 0.21 |
| Diabetes |  | 2605 (41.4%) | 527 (35.6%) | <0.001 |
| COPD |  | 1489 (23.7%) | 313 (21.1%) | 0.040 |
| Remdesivir |  | 3651 (58.0%) | 908 (61.4%) | 0.019 |
| Steroids |  | 4773 (75.8%) | 1130 (76.4%) | 0.68 |
| Hospital LOS, median (IQR) |  | 6 (3, 11) | 5 (3, 9) | <0.001 |
| ICU |  | 1435 (22.8%) | 257 (17.4%) | <0.001 |
| Mechanical Ventilation |  | 756 (12.0%) | 112 (7.6%) | <0.001 |
| Hospital Mortality |  | 1234 (19.6%) | 63 (4.3%) | <0.001 |
| Patients under age 50 |  | Nov-Jan | Mar-Apr | p-value |
|  |  | N=622 | N=283 |  |
| Age, median (IQR) |  | 40 (32, 46) | 40 (31, 45) | 0.56 |
| Race | Black | 119 (19.1%) | 80 (28.3%) | 0.002 |
|  | White | 454 (73.0%) | 191 (67.5%) |  |
|  | Other | 49 (7.9%) | 12 (4.2%) |  |
| Hypertension |  | 197 (31.7%) | 74 (26.1%) | 0.093 |
| Diabetes |  | 183 (29.4%) | 58 (20.5%) | 0.005 |
| COPD |  | 37 (5.9%) | 13 (4.6%) | 0.41 |
| Remdesivir |  | 268 (43.1%) | 154 (54.4%) | 0.002 |
| Steroid |  | 385 (61.9%) | 205 (72.4%) | 0.002 |
| Hospital LOS, median (IQR) |  | 4 (2, 8) | 4 (2, 6) | 0.15 |
| ICU |  | 141 (22.7%) | 49 (17.3%) | 0.067 |
| Mechanical Ventilation |  | 79 (12.7%) | 25 (8.8%) | 0.091 |
| Hospital Mortality |  | 28 (4.5%) | 3 (1.1%) | 0.008 |
| Abbreviations: ED=Emergency Department; COPD=Chronic Obstructive Pulmonary Disease; LOS=Length of Stay; ICU=Intensive Care Unit; IQR=Interquartile Range |  |  |  |  |
| Statistical analysis: categorical variables are displayed as N(%) and analyzed with chi-squared tests. Continuous variables are displayed as median (IQR) and analyzed using Wilcoxon Rank-Sum tests. |  |  |  |  |

Among patients under age 50, those in March and April were three times as likely to be hospitalized (12.5% vs. 4.5%, p<0.001) and twice as likely to require admission to an ICU (2.2% vs. 1.0%, p<0.001) or to need mechanical ventilation (1.1% vs. 0.6%, p=0.004) as patients under age 50 testing positive for SARS-CoV-2 from November-January (Table 1). Among hospitalized patients under age 50, those admitted in March and April were more likely to be identified as Black (28.3 vs. 19.1%, p=0.002), and rates of ICU admission were slightly lower, though the difference was not statistically significant (Table 2). More patients in March and April were treated with corticosteroids, a marker of illness severity (72.4% vs. 61.9%, p=0.002).

SARS-CoV-2 WGS was successful for three of five patients for whom it was attempted. All had infection with the alpha variant (Pangolin lineage B.1.1.7). None had completed a COVID-19 vaccine series. All were mechanically ventilated, and two died despite cannulation for extracorporeal membrane oxygenation.

## DISCUSSION

In a study of 37,502 inpatients and outpatients infected with SARS-CoV-2 from November 2020 to April 2021, we found that rates of severe illness were twice as high among younger patients during March and April compared to the peak of the pandemic from November through January, though the absolute rates remained low. Individuals identified as Black accounted for relatively more COVID-19 cases and hospitalizations over time. We identified the alpha variant in several young, previously healthy, unvaccinated patients who became severely ill in March and April. Our findings empirically validate reports that COVID-19 infections are affecting younger patients more severely than during prior periods of the pandemic, that the pandemic continues to disproportionately affect Black communities, and that novel variants may be contributing (3). These observations have significant public health implications.

Our findings underscore the continued urgency of increasing COVID-19 vaccination rates. The vaccine was not universally available until late April, which likely contributed to the relative increase in cases in younger patients in our study. The higher rate of severe disease from COVID-19 in younger patients is concerning, since rare vaccine-related adverse events may drive vaccine hesitancy despite emerging novel variants (7, 8). The vaccine timeline itself is unlikely to explain the observed increase in racial disparities—which are instead likely related to structural disparities in vaccination and continued disparities in exposure risk (2, 9). An important limitation is that these racial disparities are due to the fact that reported race is just a proxy for other socioeconomic factors which mediate racial disparities in COVID-19 and were not available in our data, rather than race itself being the causal factor (10). Our findings emphasize the need for institutions to expand intentionally-designed programs promoting access to and trust in the COVID-19 vaccine in communities experiencing disproportionate effects from the pandemic (9, 11). In the absence of sustained, effective, and equitable vaccination campaigns at local, state, and national levels, we will likely see continued deaths as well as long-term morbidity among survivors in the US throughout the remainder of 2021.

## Data Availability

Because the data come from the UPMC Health System, access to the data would require a DUA executed with the University of Pittsburgh IRB and UPMC

## ACKNOWLEDGMENTS

We thank Jane Marsh, Kady Waggle, and Marissa Griffith of the Microbial Genomic Epidemiology Laboratory, Dan Snyder of the Microbial Genome Sequencing Center, and John Mellors and Asma Naqvi of the Department of Medicine for their assistance with SARS-CoV-2 whole genome sequencing. We thank Swati Bhosale of the Department of Medicine’s Office of the Chair for assistance in data collection and serving as an honest broker. We also thank the clinical staff of the UPMC Health System for their dedicated patient care during the pandemic.

